# Somatic activating *BRAF* variants cause isolated lymphatic malformations

**DOI:** 10.1101/2021.11.03.21265682

**Authors:** Kaitlyn Zenner, Dana M. Jensen, Victoria Dmyterko, Giridhar M. Shivaram, Candace T. Myers, Cate R. Paschal, Erin R. Rudzinski, Minh-Hang M. Pham, V. Chi Cheng, Scott C. Manning, Randall A. Bly, Sheila Ganti, Jonathan A. Perkins, James T. Bennett

**Affiliations:** Seattle Children’s Hospital, Division of Pediatric Otolaryngology, Department of Otolaryngology/Head and Neck Surgery, University of Washington, Seattle, WA, 98195, USA; Vascular Anomalies Program, Seattle Children’s Hospital, Seattle, WA, 98105, USA; Center for Developmental Biology and Regenerative Medicine, Seattle Children’s Research Institute, Seattle, WA, 98101, USA; Department of Radiology, Division of Interventional Radiology, University of Washington School of Medicine, Seattle; Seattle Children’s Hospital, Department of Laboratories, Seattle, WA 98105; Center for Integrative Brain Research, Seattle Children’s Research Institute, Seattle, WA, 98101, USA; Center for Clinical and Translational Research, Seattle Children’s Research Institute, Seattle, WA, 98101, USA; Seattle Children’s Hospital, Division of Genetic Medicine, Department of Pediatrics, University of Washington, Seattle, WA, 98195, USA

## Abstract

Somatic activating variants in *PIK3CA*, the gene that encodes the p110*α* catalytic subunit of PI3K, have been previously detected in ∼80% of lymphatic malformations (LM).^1; 2^ We report the presence of somatic activating variants in *BRAF* in individuals with *PIK3CA*-negative LM. The BRAF substitution p.Val600Glu (c.1799T>A), one of the most common driver mutations in cancer, was detected in multiple individuals with LM. Histology revealed abnormal lymphatic channels with immunopositivity for BRAF^V600E^ in endothelial cells that was otherwise indistinguishable from PIK3CA positive LM. The finding that *BRAF* variants contribute to low-flow LMs increases the complexity of prior models associating low flow vascular malformations (LM and venous malformations) with mutations in the PI3K-AKT-MTOR and high flow vascular malformations (arteriovenous malformations) with mutations in the RAS-MAPK pathway.^3^ Additionally, this work highlights the importance of genetic diagnosis prior to initiating medical therapy as more studies examine therapeutics for individuals with vascular malformations.

## Main text

Disorganized morphogenesis of arteries, veins, capillaries, and lymphatic vessels results in vascular malformations, a relatively common congenital malformation associated with significant morbidity.^4^ Vascular malformations are classified into high-flow lesions, which include arteriovenous malformations (AVMs), and low-flow lesions, which include venous malformations (VeM) and lymphatic malformations (LM). Individuals with vascular malformations typically have no family history because most are caused by post-zygotic (mosaic) activating mutations in oncogenes within the PI3K-AKT and RAS-MAPK pathways.^1-3^ Treatments for vascular malformations are primarily invasive and include sclerotherapy, embolization, and open surgical resection,^4^ but the identification of specific activating mutations in well-known oncogenic signaling pathways has led to trials examining the efficacy of targeted medical therapies.^5-11^

Previous work has shown that approximately 80% of isolated LM have somatic pathogenic variants in *PIK3CA*,^1; 2; 12^ the gene that encodes for the catalytic subunit of phosphatidylinositol 3-kinase (PI3K), a component of the PI3K-AKT pathway.^13^ *PIK3CA* is the only gene associated with isolated LM to date, and the vast majority (>90%) of LM associated pathogenic variants occur at one of three locations, referred to as “hotspots”: c.1624G>A (p.E542K), c.1633G>A (p.E545K), and c.3140A>G (p.H1047R), all of which result in PI3K hyperactivation.^14; 15^ The fraction of DNA molecules that possess the pathogenic *PIK3CA* variant (referred to as the variant allele fraction or VAF) within LM tissue is typically very low (<10%),^2^ and it has been hypothesized that a fraction of LMs without a detected *PIK3CA* variant (“*PIK3CA*-negative LM”) in fact do carry a *PIK3CA* variant that was “missed” due to low level mosaicism. It is also possible that additional genes play a role. Here, we report the identification of somatic *BRAF* mutations in *PIK3CA*-negative isolated LM as well as “missed” *PIK3CA* variants in *PIK3CA*-hotspot negative LM.

LM tissue from 106 individuals was screened for the three *PIK3CA* (NM_006218.4) hotspots (p.Glu542Lys, p.Glu545Lys, and p.His1047Arg) using droplet digital polymerase chain reaction (ddPCR) assays and molecular inversion probes, as previously reported (**Supplemental Methods**).^2^ Following this screening 22 individuals remained without a detected *PIK3CA* variant. Fifteen of these individuals had sufficient DNA (14 lesion-derived and 1 cyst fluid) for further testing, which was sent for high-depth targeted sequencing using a 44 gene panel, referred to as VANseq (Vascular ANomaly sequencing, see **Supplemental Methods** and **Table S1**) throughout the rest of this paper

VANseq identified variants in 6/15 individuals (Table 1). One individual (LR18-536) had a non-hotspot *PIK3CA* variant, c.1035T>A (p.Asn345Lys) that could not have been detected by hotspot allele-specific ddPCR screening. This variant is absent from the Genome Aggregation Database (gnomAD), is predicted to be damaging by several *in silico* tools, and has been previously reported in numerous individuals with cancer as well as one individual with CLOVES syndrome.^1; 16; 17^ Functional studies have demonstrated that this substitution results in PI3K pathway hyperactivity.^14; 15^ Although not previously reported in association with isolated LM, we interpreted this variant as being pathogenic,^18^ and confirmed the presence of the variant in additional samples from that individual using ddPCR. There was variation in VAF from undetectable to 1.4% within lesion samples (**Table 1**), as we have previously described.^2^

**Table 1.**
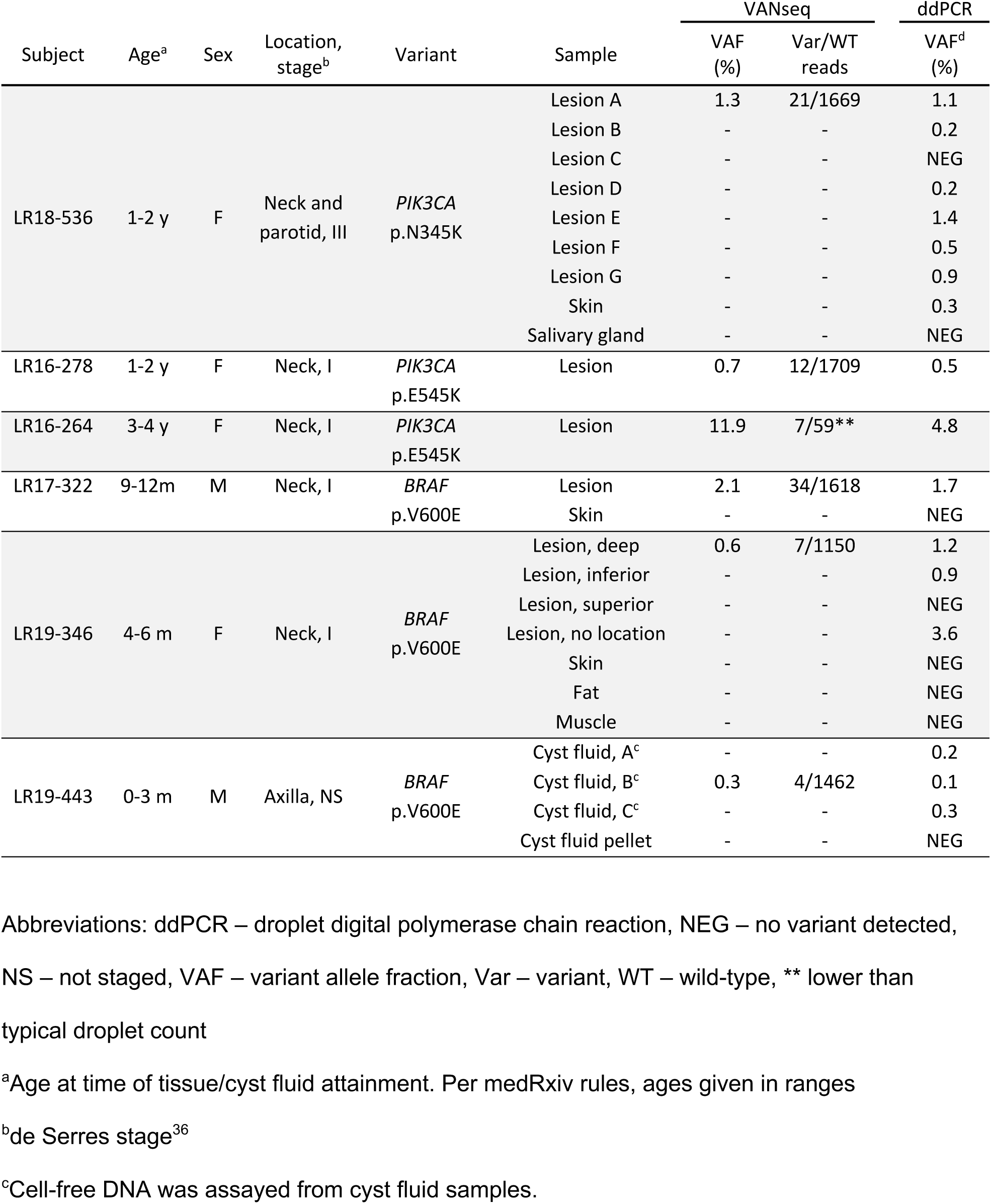
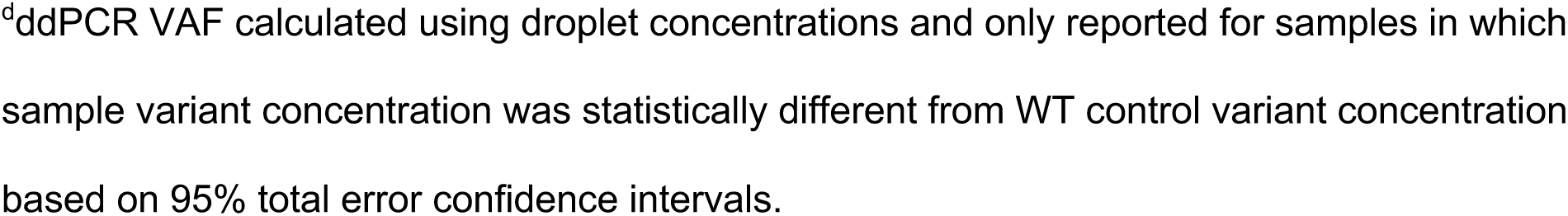
Somatic variants in LM detected by VANseq and confirmed by ddPCR.

VANseq detected a hotspot *PIK3CA* variant (p.Glu545Lys) in two individuals (LR16-278 and LR16-264), who had previously screened negative for this allele by ddPCR.^2^ We reexamined prior data from both cases. LR16-278’s prior ddPCR had 6 variant and 1055 reference droplets but did not meet our positive criteria as the 95% confidence interval overlapped with wild-type samples (**Supplemental Methods**). The initial ddPCR run for LR16-264’s had zero variant positive droplets, but subsequent testing from the same DNA isolation sample was unambiguously positive with VAF of 4.8%, possibly reflecting a sample swap during the original screening. These examples highlight difficulties in using tiered screening assays which increase the likelihood of sample swaps, and also demonstrates consideration for repeat testing when the diagnostic pre-test probability is high.^19^ We are confident that the pathogenic variant has now been identified for both individuals.

VANseq identified a pathogenic *BRAF* variant in 3 of the 15 *PIK3CA* hotspot negative individuals (LR17-322, LR19-346 and LR19-443). All three possessed the same variant (NM_004333.6:c.1799T>A, resulting in p.Val600Glu), which was confirmed by ddPCR in multiple independent tissues, when available (**Table 1, Figure 1F-H**). Two additional *PIK3CA* hotspot negative individuals (LR17-319 and LR18-572) possessed 3 or more variant reads supporting the *BRAF* p.V600E substitution but were not confirmed by ddPCR so were not classified as being *BRAF* positive (**Table S2**).

**Figure 1.**
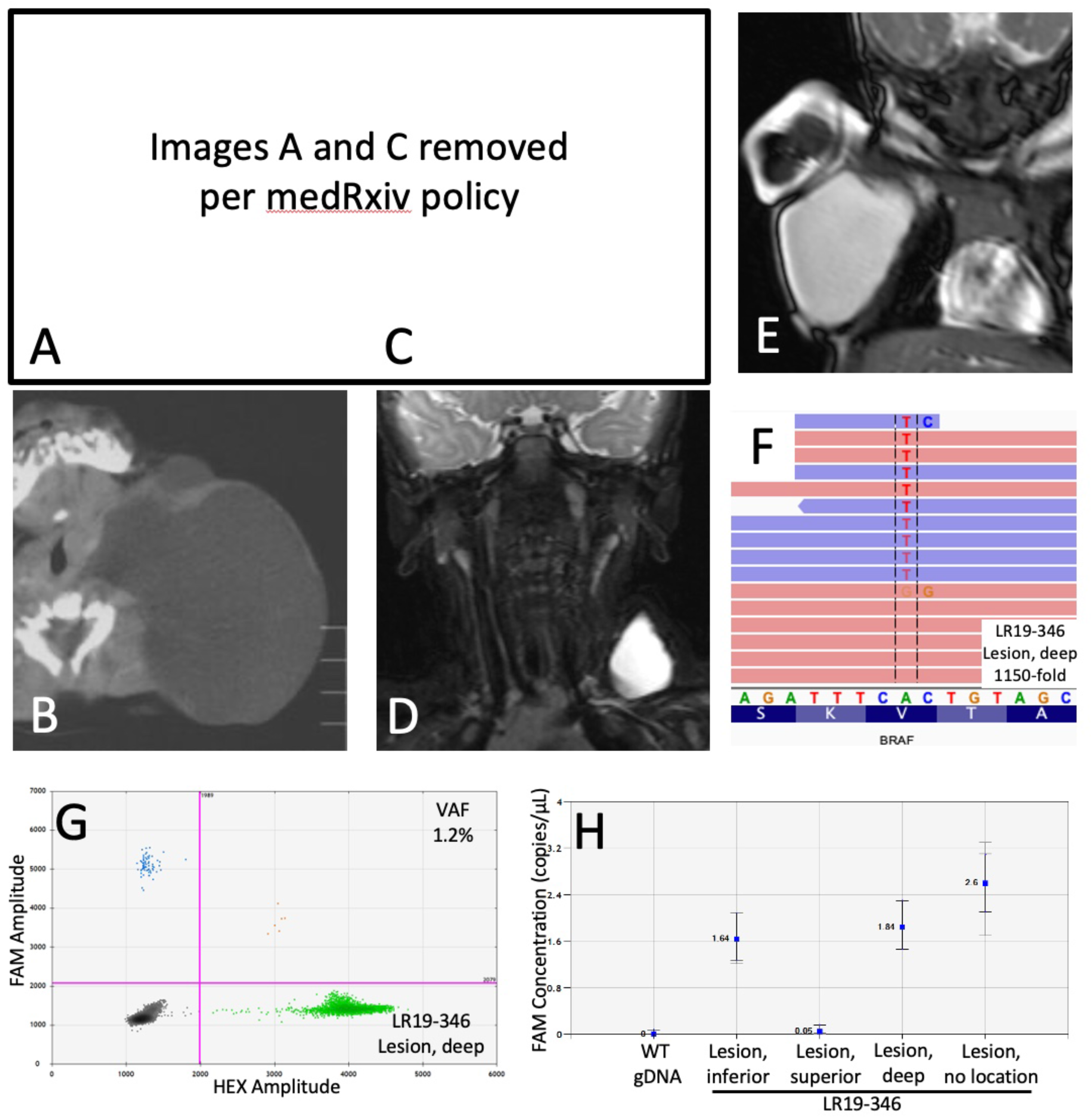
Clinical features of *BRAF*-mutated LM and confirmation of genetic diagnosis. Clinical photos of LR17-322 (A) and LR19-346 (C) showing posterior neck LM. Corresponding CT (LR17-322, B) and MRI (LR19-346, D, and LR19-443, E) images demonstrate macrocystic lesions with minimal septations of the posterior lateral neck and axilla. Integrated Genomics Viewer image for LR19-346 demonstrating somatic BRAF p.Val600Glu variant (F), confirmed on droplet digital PCR (G). Variant concentration image from Quantasoft shows variability in mutation prevalence between samples (H). *Note: Images A and B previously published prior to identification of patient’s genetic variant.^40^

All three individuals with BRAF p.V600E substitutions had macrocystic LM diagnosed at birth (**Figure 1**). LR17-322 had a large, macrocystic lesion of the posterior neck, de Serres stage 1, that resolved spontaneously over the first few months of life (**Figure 1A, B**). Surgery was performed at 1 year of age to remove remaining LM and redundant skin. LR19-346’s LM was also isolated to the neck, de Serres stage 1, and was resolving with just observation until an upper respiratory infection induced swelling and the decision was made to remove it surgically (**Figure 1C, D**). LR19-443 had a large macrocystic LM of the axilla that was treated with sclerotherapy at 1 month of age (**Figure 1E**). This individual did not have surgery, but cfDNA from aspirated cyst fluid was available for genetic diagnosis. All individuals did well after intervention with no evidence of recurrence and no further procedures or therapy.

Histopathological examination of tissues from two *BRAF* p.V600E containing LMs showed numerous dilated cystic channels with bland, flattened epithelium that was immunopositive for podoplanin, a marker of lymphatic endothelial cells (**Figure 2**).^20^ There were no distinguishing histopathological features between *BRAF* and *PIK3CA* mutant LMs. The extremely low VAFs of the BRAF p.V600E substitutions (0.3-2%) indicates that most cells within the malformation do not possess the *BRAF* substitution.^2^ We hypothesized that BRAF mutant cells would be primarily located within the lymphatic endothelial cells, as has previously been shown in *PIK3CA* positive LMs.^21-24^ To test this we used a BRAF p.V600E specific monoclonal antibody (VE1),^25^. BRAF p.V600E immunostaining was present in cyst-lining endothelial cells in LR17-322 and LR19-346 but not in other cells within the lesion (**Figure 2**). We detected no BRAF p.V600E staining in two other LM samples bearing p.E545K and p.H1047R PIK3CA substitutions (**Figure 2** and data not shown), demonstrating specificity. These results both confirm the presence of the BRAF substitutions within these lesions and demonstrate their localization to lymphatic endothelial cells.

**Figure 2.**
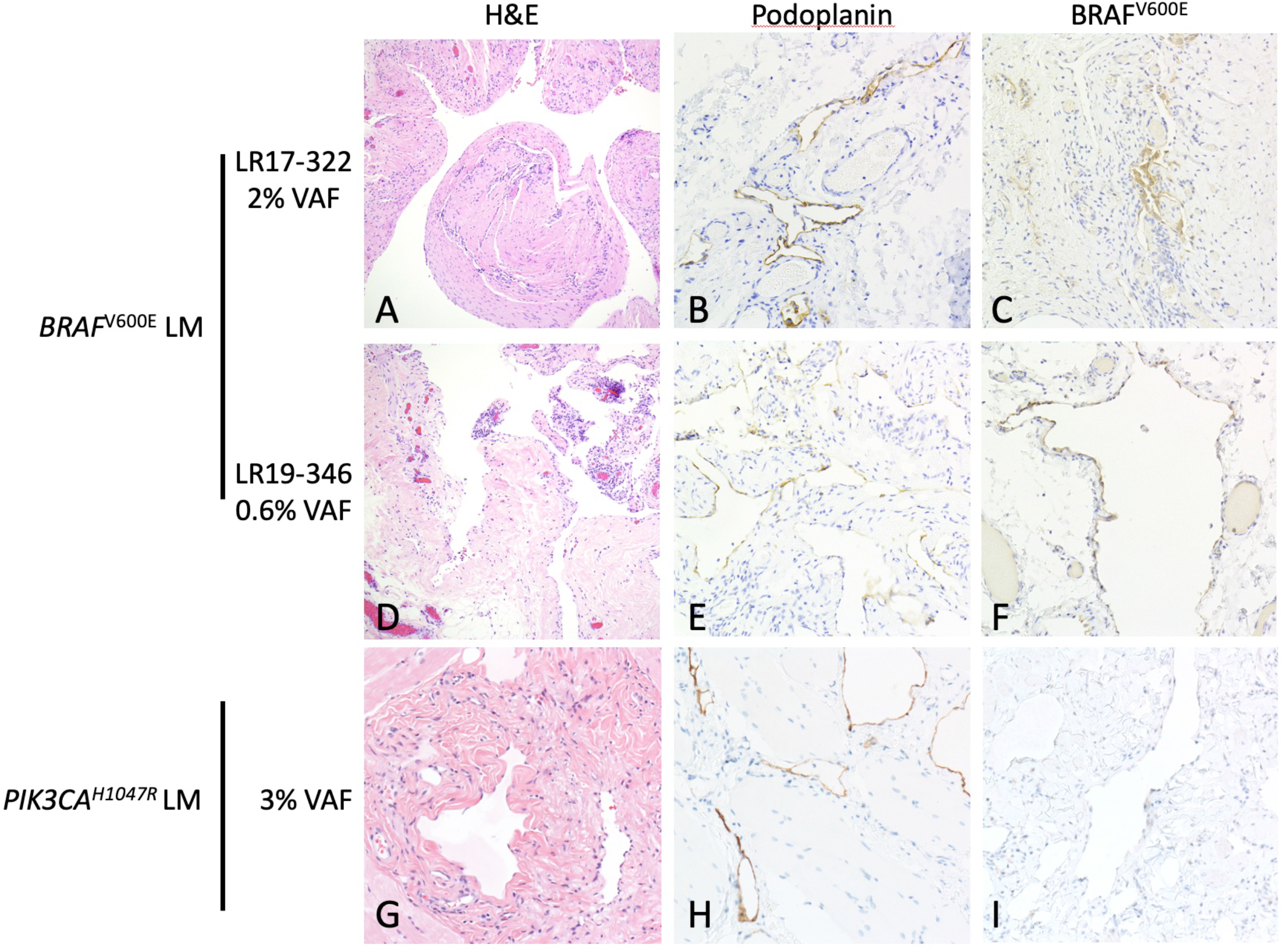
Histology and immunohistochemistry of *PIK3CA* and *BRAF* mutated LM. Lymphatic malformation tissue from two individuals with BRAF p.Val600Glu mutation (A-F) and one individual with PIK3CA p.H1047R mutation (G-I). H&E stain (panels A,D,G) show dilated cystic channels with bland, flattened epithelium. Panels B,E, and H show presence of podoplanin (aka D2-40) immunoreactivity in endothelial cells. Panels on the right shows BRAF p.V600E immunoreactivity (VE1 staining) in endothelial cells in BRAF mutant lymphatic malformation (panels C, F) but not in PIK3CA mutant lymphatic malformation (panel I).

When these results are combined with our previous reports,^2; 26^ a more complete picture of allelic and locus heterogeneity within isolated LM appears (**Table 2**). *PIK3CA* variants were found in 88% of the 101 individuals in our cohort, 92% of which occurred at one of the three *PIK3CA* hotspots. *BRAF* p.Val600Glu variants were found in 3% of individuals with isolated LM-a small proportion but a clinically important finding as responses to targeted drug therapies may differ. For example, some BRAF inhibitors produce paradoxical activation of the MAPK pathway and corresponding cellular proliferation in tumors possessing oncogenic mutations in RAS or upstream receptors.^27; 28^ The application of VANseq to our cohort of 101 individuals with isolated LM brought our overall diagnostic rate from ∼80% to over 90%, and currently only 9/101 individuals with adequate DNA now remain without a genetic diagnosis.^2^

**Table 2.** The genetic spectrum of LM including *BRAF* p.V600E.

|  | *PIK3CA* |  |  |  |  | *BRAF* |  |
| --- | --- | --- | --- | --- | --- | --- | --- |
|  | H1047R | E545K | E542K | Other | Total | V600E | NEG<sup>a</sup> |
| Zenner et al. 2019 | 22 | 18 | 18 | 6 | 64 | - | - |
| Zenner et al. 2020 | 7 | 9 | 5 | 0 | 21 | - | - |
| Current study | 1 | 2 | 0 | 1 | 4 | 3 | 9 |
| Total, N = 101<sup>b</sup> | 30 | 29 | 23 | 7 | 89 (88.1%) | 3 (3.0%) | 9 (8.9%) |

Abbreviations: NEG – no variant detected.
<sup>a</sup>All negative samples for the first two studies were included in this study if adequate sample was available for VANseq testing.
<sup>b</sup>Total includes only individuals with detected mutations or sufficient DNA to undergo VANseq testing.

*BRAF* is one of the most frequently mutated genes in cancer with a predilection for melanoma, thyroid cancer, colon cancer, and non-small cell lung cancer. p.Val600Glu is the most common oncogenic BRAF substitution, accounting for >90% of *BRAF* mutations.^29^ Non-mosaic constitutional missense and in-frame deletions in *BRAF* have been reported in rasopathies (e.g. Cardiofaciocutaneous syndrome, Noonan syndrome, and Noonan syndrome with multiple lentigines),^30^ but the p.Val600Glu substitution has never been reported in these diseases. This is likely due to the fact that the BRAF p.Val600Glu substitution is not compatible with embryonic survival except in the mosaic state (i.e,. the Happle hypothesis).^31^ This conclusion is supported by the embryonic lethality seen in constitutional expression of *BRAF* p.Val600Glu in mouse embryos.^32^

Somatic *BRAF* p.Val600Glu variants have previously been reported to cause arteriovenous malformations (AVMs), though activating mutations in *KRAS* and *MAP2K1* are more common causes.^3; 33; 34^ The precise mechanisms by which somatic BRAF p.Val600Glu substitutions cause LMs in some cases and AVMs in others likely has to do with the timing and location of the post-zygotic mutation. Additional studies are needed to examine this further. Although activating mutations in oncogenes raise concern for an increased risk of cancer, *PIK3CA*-related overgrowth syndromes have a low risk^17^ and *BRAF* p.Val600Glu variants are detected in >80% of benign melanocytic nevi, indicating that the single mutation is insufficient to produce melanoma.^35^

All three individuals with *BRAF* p.Val600Glu substitution in our study had similar clinical phenotypes – large, macrocystic lesions of the neck or body that resolved spontaneously or were treated very early in life. Under the surgical staging system for LMs (de Serres staging), these three individuals would be classified as having stage 1 lesions (unilateral and below the hyoid).^36^ Stage 1 lesions make up only ∼31% of total LM in recent studies,^2; 36; 37^, suggesting that the BRAF positive LMs may represent a milder LM phenotype than PIK3CA positive LMs. Although our cohort of BRAF positive LMs (n=3) is too small for genotype-phenotype correlations, we speculate that there may be enrichment for BRAF positive LMs among individuals with milder, non-surgical disease, as genetic diagnosis in most LMs to date has required surgically resected tissue. Non-invasive diagnostic methods such as cyst-fluid based cfDNA^26^ may provide a more balanced view of the genetic spectrum among LMs.

Endothelial cells play a key role in the pathogenesis of vascular malformations, and isolation of endothelial cells from these lesions enriches the detection of somatic variants.^21-24^ Prior work in AVM has shown *KRAS*-mutation specific staining of endothelial cells,^38^ but this not previously been possible for LM as there is no *PIK3CA*-mutant specific antibody. The presence of BRAF^V600E^ staining in lymphatic endothelial cells within the lesions supports the hypothesis that cell-non autonomous effects, such as signaling to or recruitment of wild type cells to the lesion, contribute to the formation of LMs. Cell-non autonomous effects have been previously suggested to cause cartilage overgrowth in AVMs, but additional studies will be needed to examine this further.^39^

In conclusion, we demonstrate that a somatic activating pathogenic *BRAF* variant (c.1799T>A, p.Val600Glu) is present in 3% of our cohort of individuals with isolated lymphatic malformations. Screening isolated LMs for the three PIK3CA hotspots is an efficient and cost-effective approach but will potentially miss clinically important non-hotspot *PIK3CA* and *BRAF* variation. Our use of VANseq, a high-depth, full gene sequencing panel, increased the positivity rate for our cohort of LM from ∼80 to >90%. Additionally, our results suggest the need for studies to examine the efficacy of BRAF inhibition in the treatment of lymphatic malformations.

## Data Availability

All data produced in the present study are available upon reasonable request to the authors

## Supplemental Data

Supplemental data include Supplemental Methods and Tables S1 and S2.

## Acknowledgements

We thank the participants and their families. This study was funded by the US National Institutes of Health under National Heart, Lung, and Blood Institute (NHLBI) grants F32HL147398 (to K.Z.) and R01 HL130996 (to JTB), as well as a Burroughs Wellcome Career Award for Medical Scientists 1014700 (to JTB), and a Seattle Children’s Hospital Guild Association Funding Focus Award (to JP). We also acknowledge the Seattle Children’s Vascular Anomalies Program and especially the interventional radiology team for their support in sample collection, as well as Dr. Raj Kapur for his assistance with pathology. The content of this work is solely the responsibility of the authors and does not necessarily represent the official views of the funding sources.

## Web Resources

https://cancer.sanger.ac.uk/cosmic

https://seattlechildrenslab.testcatalog.org/show/LAB1920-1

