## Supplemental material for "Somatic activating *BRAF* variants cause isolated lymphatic malformations"

**TABLE OF CONTENTS** **Page**

Supplemental Methods 2

Table S1: Gene Content of VANseq panel 4

Table S2: *BRAF* p.V600E variants on VANseq and ddPCR 4

Supplemental Citations 5

### Supplemental Methods

#### *Participants and sample collection*

This study was approved by the Institutional Review Board at Seattle Children's Hospital. Written, informed consent was obtained for each individual in this study prior to sample and data collection. All individuals presented are de-identified. We included all individuals with isolated LMs treated at Seattle Children's Hospital between 2000 and 2020 who had LM tissue available for analysis. Individuals with accompanying overgrowth syndromes such as fibro-adipose vascular anomaly (FAVA), Klippel-Trenaunay syndrome (KTS), and congenital lipomatosis, overgrowth, vascular malformations, epidermal nevi, and skeletal/spinal anomalies (CLOVES) were excluded. LM tissue was prospectively collected at clinically indicated surgical procedures, flash frozen, and stored in a biorepository at -80 degrees Celsius. DNA from lesions was isolated with PureLink Genomic DNA Mini Kit (Invitrogen, Carlsbad, CA). Blood and cyst fluid were collected in either EDTA tubes or Cell-Free DNA BCT® tubes ("Streck tubes", Streck Omaha, NE). Cyst fluid was collected during surgery or sclerotherapy, or in clinic with ultrasound guidance. cfDNA was isolated as previously described.[1] Many of the individuals included in this study were previously reported and are indicated as such in corresponding figures and tables.

#### *ddPCR screening*

Bio-Rad-designed droplet digital PCR (ddPCR) assays were used to screen the four most common *PIK3CA* mutations in LM: p.Glu542Lys, p.Glu545Lys, p.His1047Arg, and p.His1047Lys. (Bio-Rad, Hercules, CA) as previously described.[2] A subset of samples were screened for these four mutations using a *PIK3CA* multiplex ddPCR as previously reported.[1] Following identification of variants by VANseq, ddPCR assays for *PIK3CA* p.Asn345Lys and *BRAF* p.Val600Glu (Bio-Rad) were used to confirm variants and screen additional samples for variant positive individuals. Samples were positive if the variant fluorescence was significantly different from the fluorescence of the WT control using 95% confidence intervals for total error.

The total error is displayed by the QuantaSoft software and defined as the greater of either the technical error (Poisson error) or the empirical error (standard error of the mean). Variant allele fractions (VAFs) were calculated as the concentration of variant droplets out of the total concentration of droplets containing at least one copy of variant or WT DNA.

***Ultra-deep full gene sequencing: VANseq***

Target enrichment of 44 genes (**Table S1**) was performed using IDT xGen Predesigned Gene Capture Pools and custom spike-in probes. Target region includes coding exons and a minimum of 10 bp of flanking intron boundaries of the genes tested. Libraries were generated with the IDT xGen Hybridization and Wash Kit following manufacturer's instructions. Libraries were sequenced to an average depth of coverage of 1000x on a Illumina NextSeq 500 with 2x151 bp reads. Reads were aligned with Novoalign Version 2.08.02. Variants were called using samtools (mpileup) Version 0.1.19, Freebayes Version 0.9.21, GATK: Version 1.2, and Pindel: Version 0.2.4d.

***Immunohistochemistry***

Formalin fixed-paraffin embedded tissue from individuals with *PIK3CA* p.Glu545Lys and *BRAF* p.Val600Glu variants was identified in the pathology archive. Unstained slides were cut at 4 um for immunohistochemistry using the Ventana Ultra platform with the following antibodies: mouse anti-$BRAF^{V600E}$ (1:100; catalog no. ab228461; AbCam), and mouse anti-PDPN (1:10; catalog no 322M-16; Cell Marque).

**Table S1: Gene Content of VANSeq**

| | | | | |
|---|---|---|---|---|
| *ACVRL1* | *EPHB4* | *GNA14* | *MAP3K3* | *SMAD4* |
| *ARAF* | *FAT4* | *GNAQ* | *MET* | *SOX18* |
| *BRAF* | *FGFR1* | *HGF* | *NRAS* | *TEK* |
| *CCBE1* | *FLT4* | *HRAS* | *PCDC10* | *VEGFC* |
| *CCM2* | *FOXC2* | *IDH1* | *PDGFRB* | |
| *CELSR1* | *GATA2* | *IDH2* | *PIEZO1* | |
| *CTNNB1* | *GDF2* | *KIF11* | *PIK3CA* | |
| *DCHS1* | *GJC2* | *KRAS* | *PTEN* | |
| *ELMO2* | *GLMN* | *KRIT1* | *PTPN14* | |
| *ENG* | *GNA11* | *MAP2K1* | *RASA1* | |

**Table S2: *BRAF* p.V600E variants on VANseq and ddPCR**

| | VANseq | | | ddPCR | | | |
|---|---|---|---|---|---|---|---|
| Subject | Reference count | Variant count | VAF (%) | Reference count | Variant count | VAF[a] (%) | Variant detected? |
| LR17-322 | 1618 | 34 | 2.1 | 10,555 | 165 | 1.7 | Yes |
| LR19-346 | 1143 | 7 | 0.6 | 5872 | 69 | 1.2 | Yes |
| LR19-443 | 1458 | 4 | 0.3 | 9661 | 12 | 0.1 | Yes |
| LR17-134 | 1507 | 4 | 0.3 | 7756 | 0 | NEG | No |
| LR17-319 | 1256 | 4 | 0.3 | 10,850 | 0 | NEG | No |
| LR18-572 | 1420 | 3 | 0.2 | 10,558 | 0 | NEG | No |

Abbreviations: ddPCR – droplet digital polymerase chain reaction, NEG – no variant detected, VAF – variant allele fraction

[a]ddPCR VAF calculated using droplet concentrations and only reported for samples in which sample variant concentration was statistically different from wild-type control variant concentration based on 95% total error confidence intervals.
